# Causal relationships between genetically predicted circulating levels of amino acids and non-alcoholic fatty liver disease risk: a Mendelian randomisation study

**DOI:** 10.1101/2023.02.03.23285451

**Authors:** Jian Zhao, Jing Zeng, Dong Liu, Jun Zhang, Fei Li, Giovanni Targher, Jian-Gao Fan

## Abstract

**Background:** Emerging metabolomics-based studies suggested links between amino acids metabolism and non-alcoholic fatty liver disease (NAFLD) risk, however, whether there exists an aetiological role of amino acid metabolism in NAFLD development remains unknown. The aim of the present study was to assess the causal relationship between circulating levels of amino acids and NAFLD risk.

**Methods:** We performed two-sample Mendelian randomisation (MR) analyses using summary level data from genome-wide association studies (GWAS) to assess causal relationships between genetically predicted circulating levels of amino acids and NAFLD risk. Data from the largest GWAS on NAFLD (8,434 cases and 770,180 controls) were used in discovery MR analysis, and from a GWAS on NAFLD (1,483 cases and 17,781 controls) where NAFLD cases were diagnosed using liver biopsy, were used in replication MR analysis. Wald ratios or multiplicative random-effect inverse variance weighted (IVW) methods were used in the main MR analysis, and weighted median and MR-Egger regression analysis were used in sensitivity analyses. We additionally performed an MR conservative analysis by restricting genetic instruments to those directly involved in amino acid metabolism pathways.

**Findings:** We found that genetically predicted higher alanine (OR=1.45, 95% CI 1.15-1.83) and lower glutamine (OR = 0.81, 95% CI 0.66-1.00) levels were associated with a higher risk of developing NAFLD. Results from MR sensitivity analyses and conservative analysis supported the main findings.

**Interpretation:** Genetically predicted higher circulating levels of alanine was associated with an increased risk of NAFLD, whereas higher glutamine was associated with a decreased risk of NAFLD.

**Funding:** This work was supported by Xinhua Hospital, Shanghai Jiao Tong University School of Medicine (2021YJRC02).

**Research in context:** *Evidence before this study:* Recent metabolomics studies revealed associations between circulating levels of several amino acids and non-alcoholic fatty liver disease (NAFLD) risk. Most of these studies were conducted with a focus on the profiling of amino acids between individuals with NAFLD and healthy subjects, which suggested the altered amino acid metabolism might be a consequence of NAFLD rather than a causal risk factor for NAFLD. We searched PubMed for studies in any language using the search terms “amino acids” AND “Non-alcoholic fatty liver disease OR NAFLD OR fatty liver” AND “Mendelian randomisation OR Mendelian randomization”, and found few studies on the causal effects of circulating amino acids on NAFLD risk. Thus, whether there is an aetiological role of amino acids in NAFLD development remains unknown.

*Added value of this study:* In the present study, we systematically investigated the causal effects of genetically predicted circulating levels of 20 amino acids on NAFLD risk using data from large-scale genome-wide association studies in up to 778,614 individuals of European ancestry. We utilised a state-of-art causal inference approach, that is Mendelian randomisation, to construct layers of evidence. Overall, we found that among 20 amino acids, genetically predicted higher circulating levels of alanine was associated with an increased risk of NAFLD, whereas higher glutamine was associated with a decreased risk of NAFLD.

*Implications of all the available evidence:* Our study is the first to systematically assess the causal relationships between levels of plasma amino acids and the development of NAFLD using multi-omics (i.e., genomic and metabolomic) data from large-scale human studies. Our results suggest the potential for the glutamine supplementation or alanine depletion for personalized nutrition in NAFLD prevention and treatment.

## Introduction

Non-alcoholic fatty liver disease (NAFLD) is one of the most prevalent chronic liver diseases, affecting up to ∼30% of the general population globally ^1^. NAFLD has been also predicted to become the most frequent indication for liver transplantation in Western countries by 2030 ^2^. NAFLD is a progressive disease characterized by the accumulation of lipid droplets within hepatocytes in the absence of excessive alcohol consumption and defined by the presence of at least 5% hepatic steatosis ^3^. This condition has been consistently reported to be associated with important cardiometabolic comorbidities, including obesity, type 2 diabetes mellitus, cardiovascular disease and stroke ^4,5^. To date, although substantial efforts have been put forth to prevent or treat NAFLD, there are no effective preventions or therapeutic treatments for NAFLD.

Emerging metabolomics-based studies have provided insights into mechanisms underlying the development and progression of NAFLD ^6,7^. Identifying pathogenic molecules of NAFLD development is essential for improving aetiological understanding and developing novel therapeutic targets for early intervention of this common and burdensome liver disease. It is known that abnormal lipid and glucose metabolism exert putative roles in the pathogenesis of NAFLD ^8^, whereas recent studies suggested that amino acid metabolism might also contribute to the pathogenesis of NAFLD ^9,10^. For example, lower glycine was reported to be associated with higher prevalence of NAFLD ^11^. Increased levels of aromatic amino acids (AAAs) (e.g., tyrosine and phenylalanine) were found to be associated with increased risk of liver diseases ^12^. Increased levels of branched chain amino acids (BCAAs), including leucine, isoleucine and valine, have also been reported during the progression of NAFLD ^13^. In addition, a recent Mendelian randomisation study found a causal effect of NAFLD on blood tyrosine levels ^14^. Most previous studies have focused on the profiling of amino acids or altered amino acid metabolism in individuals with NAFLD, compared to those without NAFLD, for discovery of non-invasive diagnostic biomarkers. Metabolism of amino acids including BCAAs, alanine, glutamine and tyrosine has been reported to be impacted by NAFLD ^13,15^. This implies that altered metabolism of amino acids might be a consequence of NAFLD rather than a causal risk factor for NAFLD. Thus, whether there exists an aetiological role of amino acid metabolism in NAFLD development (i.e., a causal effect of circulating levels of amino acids on NAFLD risk) remains currently unknown.

Mendelian randomisation (MR) is a causal inference approach using germline genetic variants as instrumental variables (IVs), which could largely minimize the risk of bias due to residual confounding or reverse causation ^16,17^. In this study, we implement two-sample MR analyses to systematically assess causal effects of genetically predicted circulating levels of amino acids on risk of NAFLD using summary data from both the latest and largest genome-wide association studies (GWASs) of human metabolites and two independent GWASs of NAFLD (**Figure 1 Panel A**).

**Figure 1.**
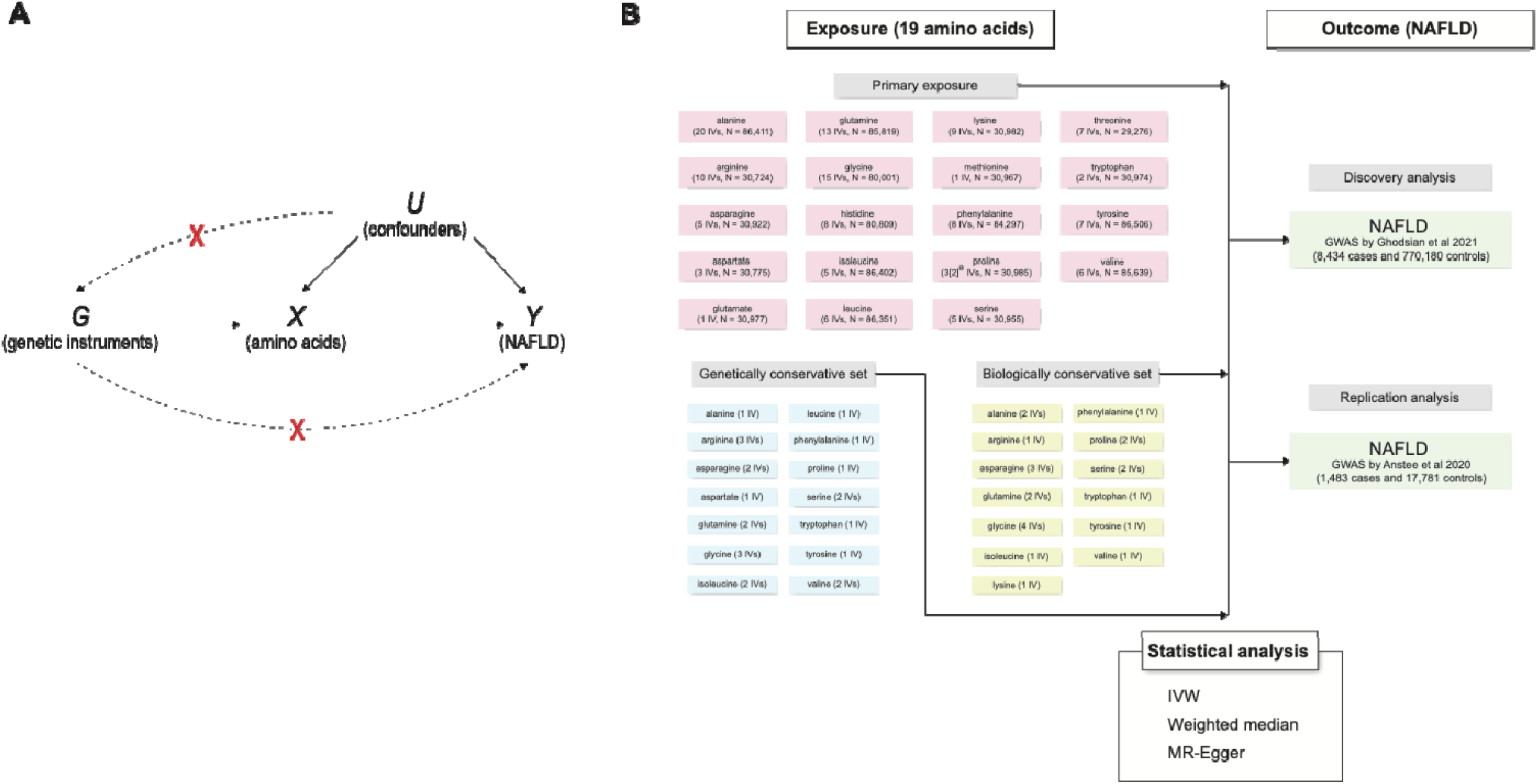
Schematic overview of the study design and MR analysis. a. rs3970551 was absent from the IV set in the replication NAFLD GWAS by Anstee et al. due to non-available proxy SNPs being identified.

## Methods

### Data sources

#### Exposure measure: Amino acids

Summary data for genetic associations with amino acids were retrieved from a recently conducted cross-platform GWAS of 174 metabolites that included up to 86,507 participants (for individual metabolites sample sizes varied from 8,569 to 86,507) ^18^. Genome-wide association results were meta-analysed in three cohort studies (i.e., the Fenland, EPIC-Norfolk and INTERVAL studies) followed by a further meta-analysis with publicly available GWAS summary data from two studies ^19,20^. Of 174 plasma metabolites investigated in the GWAS, 20 circulating levels of amino acids (alanine, arginine, asparagine, aspartate, cysteine, glutamate, glutamine, glycine, histidine, isoleucine, leucine, lysine, methionine, phenylalanine, proline, serine, threonine, tryptophan, tyrosine and valine) were included. We calculated the SNP specific genetic associations with amino acids and their corresponding standard errors based on the original information, including the z-score, sample size and minor allele frequency (MAF), reported in the GWAS of metabolites using the following formula 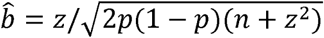 and 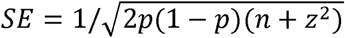 ^21^,where b is the SNP specific genetic association, z is the z-score, p is the MAF of the SNP, n is the sample size, and SE is the standard error of the genetic association.

### Outcome measure: NAFLD

Two independent datasets on the outcome measure (i.e., NAFLD) were namely retrieved from recently conducted GWASs of NAFLD ^22,23^. Data for discovery analysis were obtained from the largest NAFLD GWAS meta-analysis conducted in four cohorts of European ancestry: the Electronic Medical Records and Genomics (eMERGE) network, the UK Biobank (UKB), the Estonian Biobank (EstBB) and the FinnGen ^23^. In the GWAS meta-analysis on NAFLD, two GWASs of NAFLD were firstly conducted in the UKB and EstBB cohorts, and then combined with results from two publicly available NAFLD GWASs (eMERGE and FinnGen). As a result, 8,434 NAFLD cases were identified by electronic health records (EHR) and 770,180 controls were included in the GWAS meta-analysis (**Figure 1 Panel B**). Furthermore, for the replication analysis, data were retrieved from a large GWAS of NAFLD in individuals of European ancestry (1,483 cases and 17,781 controls), where NAFLD cases were diagnosed using liver biopsy (i.e., the gold standard method for diagnosing NAFLD) ^22^ (**Figure 1 Panel B**).

### Genetic instruments selection and data harmonization

Based on the GWAS summary data on cross-platform measured metabolites, 112 genetic variants, which were associated with at least one of 20 circulating amino acids at a metabolome-adjusted genome-wide significance level (p < 5 × 10^−8^ / 102 = 4.9 × 10^−10^), were selected as candidate instrument variables (IV). In the present study, a stringent linkage disequilibrium (LD) clumping threshold (r2 < 0.01 and window = 10 Mb) for genetic instruments selection was applied using the “clump_data” function in the TwoSampleMR R package. A total of 111 SNPs (excluding rs61937878) were retained after LD clumping.

Each genetic instrument was looked up in two NAFLD GWASs (for discovery and replication analysis respectively) for SNP-NAFLD associations. SNPs that are in high LD with genetic instruments (r^2^ > 0.8 and window = 500 Mb) were identified to proxy the absent variants in the discovery and replication NAFLD datasets (14 and 6 proxies were identified respectively). Detailed information on the proxy SNPs can be found in the **Supplementary Table S1**. Two SNPs (rs142714816, the unique instrument for cysteine, and rs3970551), both of which were absent from the summary data of NAFLD GWAS with no proxy SNPs available, were excluded from the discovery analysis, and one SNP (rs142714816) was removed from the replication analysis.

A data harmonization procedure was performed to merge SNP-amino acid and SNP-NAFLD associations using the “harmonise_data” function in the TwoSampleMR R package ^24^. Two palindromic SNPs (rs2422358 and rs1935) were removed from further analysis. As a result, a total of 107 and 108 eligible SNPs used as instrumental variables for 19 circulating amino acids were included in the discovery and replication MR analysis, respectively.

### Statistical analysis

In both discovery and replication stages, we used Wald ratios (for glutamate and methionine because only one SNP was available for each of these two amino acids), and multiplicative random-effect inverse variance weighted (IVW) approach for all other amino acids as the main MR analysis method, to estimate the causal effect of genetically predicted circulating levels of amino acids on risk of having NAFLD. To increase the statistical power and precision of the causal estimates, a fixed-effect meta-analysis was performed to combine the causal estimates in both discovery and replication stages using the meta R package ^25^.

Additionally, for certain amino acids that have five or more genetic instruments, we performed several sensitivity analyses, including weighted median and MR-Egger regression analysis to test the consistency of the causal estimates under the different assumptions and to detect possible pleiotropy. Unlike the IVW method that assumes all the SNPs are valid IVs ^26^, the MR-Egger regression could generate a consistent estimate in presence of invalid genetic instruments, as long as the Instrument Strength Independent of Direct Effect (InSIDE) assumption holds ^27^. The weighted median method assumes that more than half of the genetic instruments are valid and is a robust approach to outliers ^28^.

To assess the strength of the selected genetic instruments in MR analysis, we calculated F statistic for each genetic instrument, which are generally considered strong when greater than 10 ^29^. We used Cochrane’s Q statistic to examine the heterogeneity between SNP-specific causal estimates. Substantial heterogeneity between SNP-specific causal estimates could be indicative of horizontal pleiotropy.

Furthermore, to minimize the risk of bias due to horizontal pleiotropy, we also performed a conservative MR analysis by restricting genetic instruments to those directly involved in amino acid metabolism pathways, as described elsewhere ^30^. Two sets of genetic variants, namely biologically and genetically prioritised conservative SNPs, were used as instrumental variables in conservative MR analysis to estimate causal effects using the Wald ratios or fixed-effect IVW method as appropriate.

Given that a total of 20 amino acids were investigated in the present study, after a multiple testing Bonferroni correction, an estimate with a p-value <0.0025 (p=0.05/20) was considered as strong evidence for causal effects, whereas a p-value between 0.0025 and 0.05 indicated a suggestive causal effect. All statistical analyses were undertaken with R version 4.0.2 (R Foundation for Statistical Computing, Vienna, Austria).

### Ethics statement

Ethics approval has been obtained in original studies that contributed to GWASs on amino acids and NAFLD. All participants provided written informed consent. Declaration of Helsinki statement has been described in the original publications of these studies. The present study only used summary level data from relevant GWASs.

## Results

### Characteristics of the included studies and the selected SNPs

Genetic variants instrumenting for amino acids in our study were obtained from a meta-analysis of metabolites GWAS using data from up to 23 cohorts included in previous three GWASs ^20,31,32^ and three independent studies (the Fenland cohort ^33^, EPIC-Norfolk Study ^34^, and INTERVAL trial ^35^) (**Table 1**). Average participant age of included studies ranged from 43.5 to 59.8 years old ^36^. Approximately 50.4% to 53.9% of the study participants were women, except for the GWAS conducted by Shin et al.,^20^ where only 16.5% of participants were women. A large-scale meta-analysis of NAFLD GWASs in four studies of European ancestry (the eMERGE, FinnGen, UKB, and EstBB cohorts) included 8,434 NAFLD cases and 770,180 controls and were used for discovery MR analysis ^23^. Another independent NAFLD GWAS used for the replication analysis included 1,483 NAFLD cases diagnosed with liver biopsy and 47.3% of these participants were women ^22^.

**Table 1.**
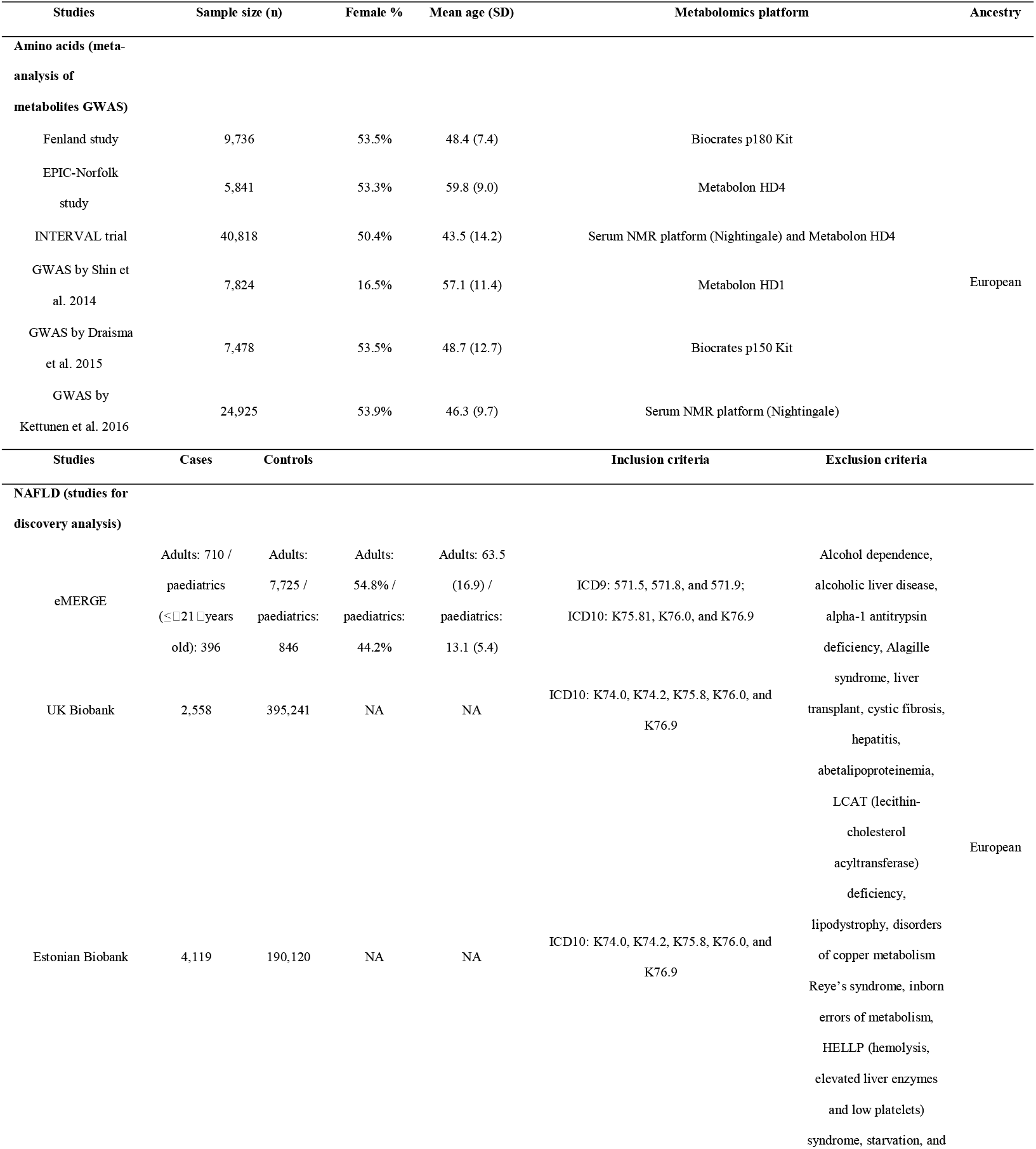
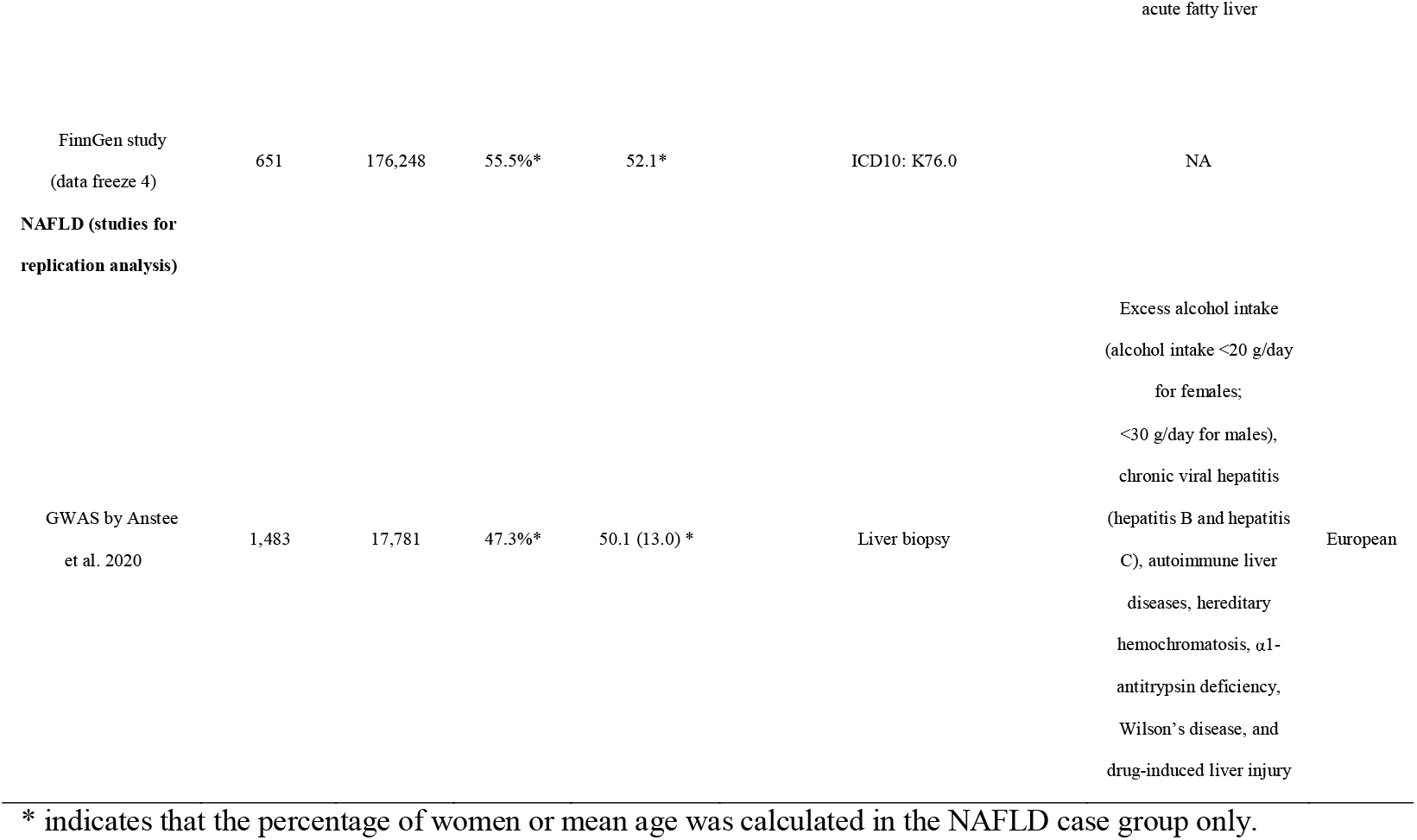
Characteristics of included studies.

The characteristics of the selected SNPs instrumenting for amino acids are presented in **Supplementary Table S2**. A total of 133 and 134 genetic variants were used as IVs, ranging from 1 IV (for glutamate and methionine) to 20 IVs (for alanine), to estimate the causal effects of 19 amino acids on NAFLD in discovery and replication MR analysis, respectively. The F-statistics of genetic variants instrumenting for 19 amino acids ranged from 38.7 to 7504.1 (**Supplementary Table S2**), suggesting a low risk of weak instrument bias. Proportions of variation in amino acids explained by genetic instruments ranged from 0.13% (glutamate) to 10.38% (glycine) (**Supplementary Table S3**). Cochrane’s Q statistics indicated that there was no significant heterogeneity between SNP-specific causal estimates for arginine, aspartate, phenylalanine, proline, and tryptophan in the discovery MR analysis (**Supplementary Table S4**).

### MR main analysis results

Of 19 amino acids examined, genetically predicted higher circulating alanine levels were causally associated with an increased risk of NAFLD in both discovery and replication analyses. The odds ratio (OR) of NAFLD was 1.45 (95% CI 1.15-1.83; p = 0.002) per 1-SD increment in alanine levels, after combining causal effect estimates from discovery (OR = 1.37, 95% CI 1.07-1.76; p = 0.012) and replication (OR = 2.06, 95% CI 1.08-3.94; p = 0.029) MR analyses (**Figure 2**). Additionally, genetically predicted higher circulating glutamine levels appeared to be suggestively associated with a lower risk of NAFLD (OR = 0.81, 95% CI 0.66-1.00; p = 0.048) after meta-analysing estimates from discovery (OR = 0.80, 95% CI 0.64-1.02; p = 0.068) and replication (OR = 0.84, 95% CI 0.55-1.29; p = 0.436) MR analyses (**Figure 2**). There was little evidence for a causal association between circulating levels of the remaining amino acids and NAFLD risk. Causal effect estimates from the replication MR analysis were broadly consistent with that from the discovery analysis, except for methionine, which had discrepant directions of effect but low precisions.

**Figure 2.**
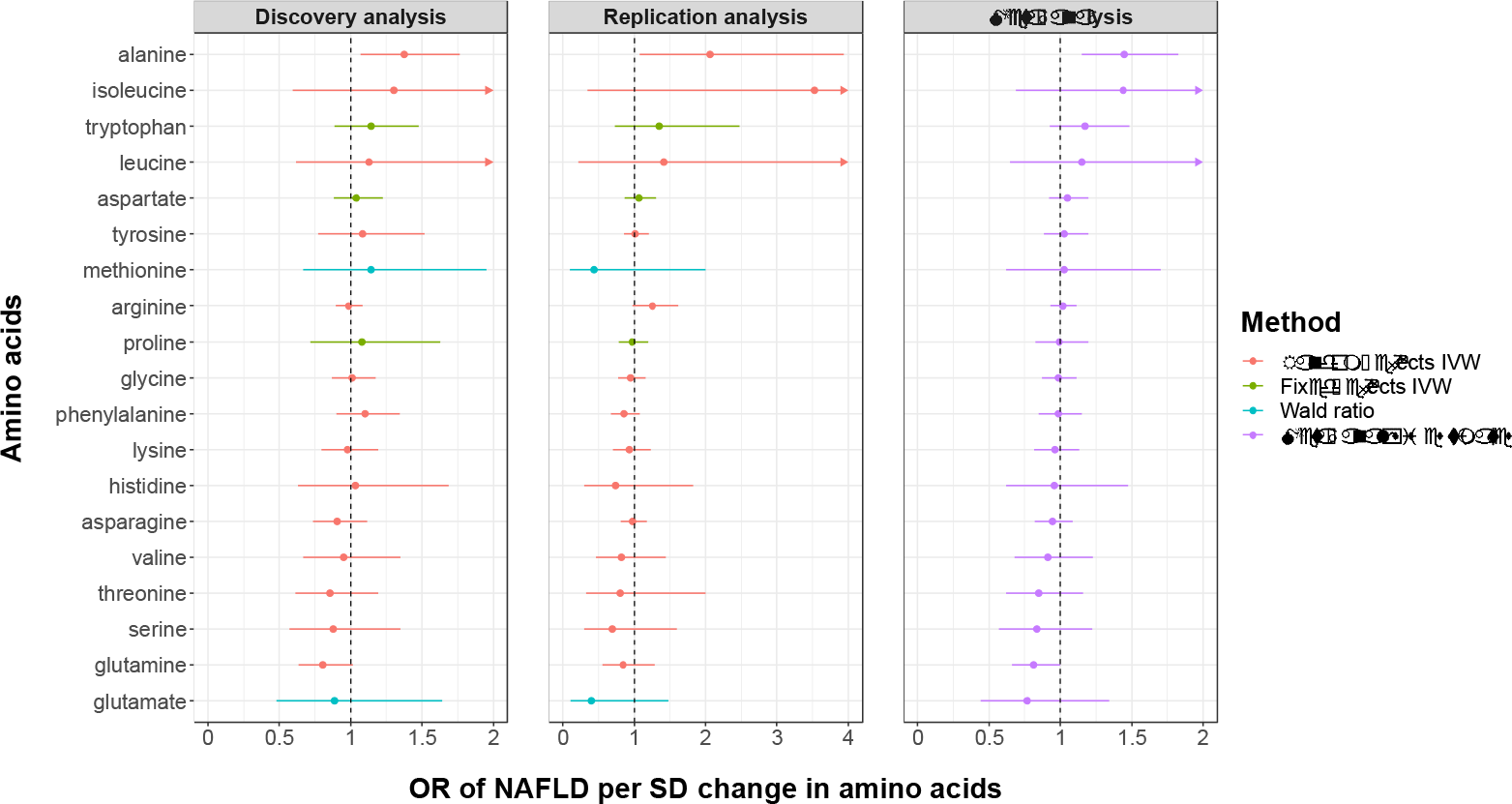
MR main analysis results of the causal effects of genetically predicted circulating levels of amino acids on NAFLD risk.

### MR sensitivity analyses results

MR sensitivity analyses, including weighted median and MR-Egger regression analyses, were conducted in 14 amino acids that had at least 5 SNPs as genetic instruments (**Supplementary Table S5**). A broad consistence was observed when comparing results from sensitivity analyses with those from main analysis presented above, except for several amino acids which had very low precisions in replication MR-Egger regression analysis. Reasons for the low precisions of estimates included smaller sample size of data used in replication analysis and homogeneous SNP-amino acid associations ^37^. Of note, meta-analysed causal effect estimates derived from weighted median analysis, which is statistically more robust compared to MR-Egger regression, supported potential causal effects of both alanine (OR = 1.58, 95% CI 1.22, 2.04; p < 0.001) and glutamine (OR = 0.81, 95% CI 0.70, 0.93; p = 0.004) on NAFLD risk.

### Conservative MR analysis results

By restricting genetic instruments for amino acids to SNPs that were biologically or genetically prioritized in previous published GWAS of metabolites ^18^, we performed a conservative MR analysis to achieve more reliable causal inference. We were unable to investigate histidine, threonine, methionine and glutamate as genetic variants instrumenting for these amino acids were not on the list of biologically or genetically prioritized genes nor directly involved in relevant metabolism pathway. Results from the conservative MR analysis confirmed a causal role of alanine (OR = 1.80, 95% CI 1.09, 2.97, p = 0.022 for biologically prioritised IVs; OR = 1.93, 95% CI 1.26, 2.96, p = 0.003 for genetically prioritised IVs) and glutamine (OR = 0.83, 95% CI 0.73, 0.96, p = 0.009 for both biologically and genetically prioritised IVs) on the risk of NAFLD (**Figure 3**).

**Figure 3.**
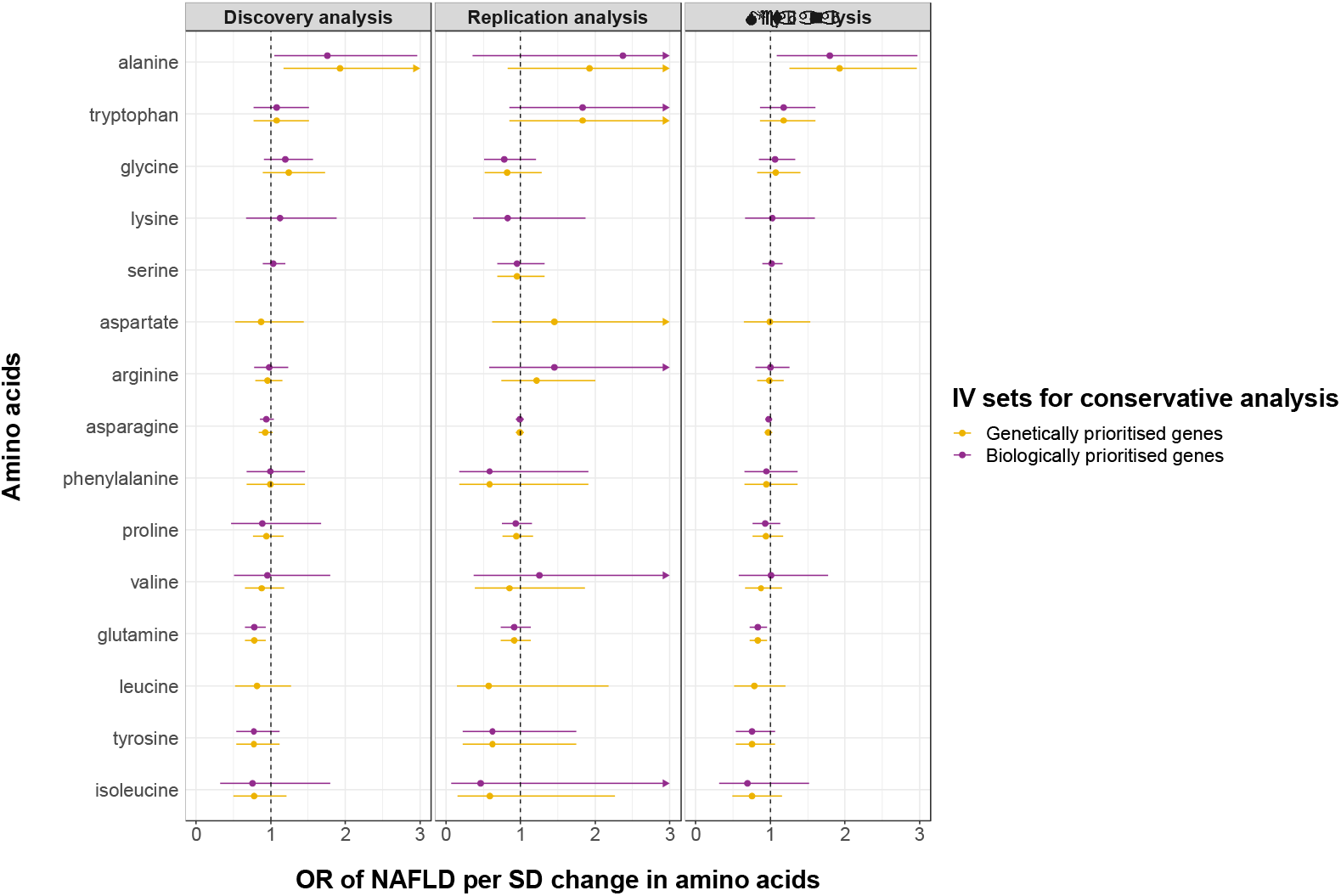
MR conservative analysis results using genetically and biologically prioritised variants as instrumental variables.

## Discussion

In this MR study, we provided novel evidence for a causal role of genetically predicted circulating levels of alanine and glutamine in the development of NAFLD. Specifically, genetically predicted higher alanine and lower glutamine were associated with a higher risk of developing NAFLD. To the best of our knowledge, it is the first study systematically assessing the causal relationships between levels of plasma amino acids and the development of NAFLD using multi-omics (i.e., genomic and metabolomic) data from large-scale human studies (in up to 778,614 individuals).

Previous observational studies mainly focused on the profiling of amino acids or altered amino acid metabolism in individuals with NAFLD compared with those without NAFLD. Metabolism of amino acids including BCAAs (i.e., leucine, isoleucine and valine), alanine, glutamine and tyrosine has been reported to be impacted by NAFLD ^13,15^. These findings were beneficial to identifying diagnostic biomarkers of NAFLD, whereas they were not capable to provide causal evidence for aetiological biomarkers of NAFLD development. Thus, our findings provide novel insights into the causal mechanism between altered amino acid metabolism and NAFLD development.

Glutamate is one of the major substrates for the synthesis of glutathione, which is a tripeptide consisting of glutamate, cysteine and glycine and protects tissues from free radical injury via detoxification of active species and/or repair of injury. Since glutamate is poorly transported into cells and glutamine can be efficiently transported across the cell membrane and deaminated in the mitochondria to produce glutamate and NH3, plasma glutamine is thus important for the generation of intracellular glutamate and consequently glutathione.

Experimentally, it has been reported a potential causal role of glutamine administration in decreasing liver injury and mortality in animal studies ^38-40^. However, there are sparce human studies on the effect of glutamine administration on liver function and its related biomarkers. The present study, from a genetic perspective, provides causal evidence for a protective causal effect of higher circulating glutamine levels on the development of NAFLD. Further, in our MR conservative analysis, we found that only the *GLS-2* (rs2657879) genetic variant predicted glutamine exerting a causal effect on NAFLD risk, compared with another variant (*GLS*, rs7587672) instrumenting for glutamine. Our results were partly supported by findings from a previous study, where the authors found that reducing glutamine metabolism (loss-of-function of *GLS2)* in the liver resulted in decreased severity of hyperglycaemia (increased plasma levels of glutamine and reduced levels of fasting glucose) ^41^.

Alanine is the predominant amino acid contributing to hepatic gluconeogenesis, therefore, abnormal levels of which generally indicate dysregulation of the alanine-glucose cycle, even further consequent tricarboxylic acid (TCA) and urea cycles ^42,43^. Elevated alanine concentrations in NAFLD have been observed in multiple latest metabolomics studies ^44^, which was consistent with an emerging hypothesis of dysregulated TCA and urea cycles in NAFLD ^45,46^. Recently, in the Young Finns Study that examined prospective associations between baseline metabolite levels and the future risk of NAFLD, plasma alanine levels were also found to be positively associated with future onset of NAFLD ^47^. In our study, results from MR analysis confirmed the positive causal effect of circulating levels of alanine on NAFLD risk.

Among other amino acids, higher levels of BCAAs (including leucine, isoleucine and valine) and AAAs (including phenylalanine, tryptophan, and tyrosine) have been reportedly linked with NAFLD ^48-50^, however, to our knowledge, we identified only one study in which the prospective associations between baseline concentrations of amino acids and the risk of developing NAFLD during 10-year follow-up were examined ^47^. Interestingly, in our study, on contrary to the positive associations revealed in the above-mentioned studies, we find little evidence to support a causal effect of both BCAAs and AAAs on NAFLD development. One reason for these discordant results might be reverse causation or confounding bias that cannot be ruled out in previous observational studies. For example, in the prospective Young Finns Study, increased plasma tyrosine levels were associated with higher 10-year risk for fatty liver when first adjusted for sex and age, whereas after adjusting for additional baseline confounders, such as waist circumference, alcohol intake, smoking and leisure-time physical activity, this association attenuated and became statistically non-significant ^47^. Further, in a recent MR study investigating the causal effect of NAFLD on consequent blood metabolites, NAFLD was found to have a positive impact on plasma tyrosine levels ^14^. Taken together with our results, it seems more plausible to consider altered tyrosine metabolism as a response to the presence of NAFLD rather than an aetiological factor for NAFLD development.

Our study has several strengths. Firstly, this is the first and largest study systematically investigating the causal effects of human circulating amino acids on NAFLD risk, utilizing multi-omics data. Secondly, we leveraged data from an independent GWAS of NAFLD to validate our findings in the discovery population, and combined causal effect estimates from both datasets using meta-analysis to increase statistical power and estimate precision. Thirdly, the conservative MR analysis that was less susceptible to horizontal pleiotropy using genetically and biologically prioritized SNPs as instrumental variables confirmed findings from our MR main analysis. Finally, our results can be generalized to European ancestry as samples span the entire Europe.

We acknowledge some important limitations of our study. Firstly, our study was limited to individuals of European ancestry due to data availability, thus generalizability to other ethnic populations needs to be further investigated. Secondly, although summary data from the largest histology based NAFLD GWAS was used to replicate our findings, results derived from discovery analysis were based on electronic health record (EHR) data where diagnosis of NAFLD may be biased by misclassification of cases and controls due to using hospital records (i.e., ICD-9 and ICD-10 codes). Therefore, future replications in larger cohorts of participants with NAFLD diagnosed with gold standard (i.e., liver biopsy) are warranted.

In conclusion, novel causal biomarkers including alanine and glutamine of NAFLD development were revealed in our study with integrating genomic and metabolomic data. Although further studies are needed, these findings suggest the potential for the glutamine supplementation or alanine depletion for personalized nutrition in NAFLD prevention and treatment.

## Data Availability

Genetic association estimates for amino acids were obtained by accessing to omicscience web (https://omicscience.org). The summary statistics on NAFLD were obtained from GWASs conducted by Ghodsian et al. and Anstee et al., which had been deposited at the GWAS catalog (https://www.ebi.ac.uk/gwas/). 

## Contributors

Jian Zhao: Study conception and design; data curation and analysis; drafting and reviewing the article. Jing Zeng: Data curation; drafting and reviewing the article. Dong Liu: Data curation and analysis; reviewing the article. Jun Zhang: Reviewing the article. Fei Li: Reviewing the article. Giovanni Targher: Drafting and reviewing the article.: Jian-Gao Fan: Study conception and design; data curation; drafting and reviewing the article.

## Data sharing statement

Genetic association estimates for amino acids were obtained by accessing to ‘omicscience’ web (https://omicscience.org). The summary statistics on NAFLD were obtained from GWASs conducted by Ghodsian et al. and Anstee et al., which had been deposited at the GWAS catalog (https://www.ebi.ac.uk/gwas/).

## Acknowledgements

This study was supported by Xinhua Hospital, Shanghai Jiao Tong University School of Medicine (2021YJRC02). The authors would like to thank all GWASs investigators for sharing these valuable data.

